# Neural aperiodic activity as a novel test of vigilance: a proof-of concept, retrospective study of patients undergoing MWT

**DOI:** 10.64898/2025.12.16.25342307

**Authors:** Kevin Seow, Michelle Bay, Samuel Brookes, Teng Y Kang, Sonya Johnston, Aeneas Yeo, Kurt Lushington, Alex Chatburn

## Abstract

**Background:** The maintenance of wakefulness test (MWT) measures the ability of an individual to maintain wakefulness in soporific conditions and despite limitations remains the mainstay of vigilance testing. Wake and sleep states are traditionally characterised by oscillatory activity on electroencephalogram (EEG) but there is a less utilised non-oscillatory component of background neural aperiodic activity which can be derived from EEG raw data and represented as an exponent or a gradient (1/*f* slope). There is emerging evidence that aperiodic activity can be used to predict cortical activity. The aim of this study is to assess the use of aperiodic activity as a novel neurobiological marker of vigilance.

**Methods:** EEGs (4 x 50) from 50 patients who underwent the MWT between 2009 and 2023 at a single centre were analysed to determine their aperiodic activity. Statistical analyses utilising linear mixed models and linear regression were performed to assess the relationship between aperiodic exponent, aperiodic intercept and mean sleep latency.

**Results:** Linear mixed-effects modelling revealed that more negative aperiodic exponents represented by steeper 1/*f* slopes were associated with longer sleep onset latencies (β=-8.08, *p*=0.002). Similarly, higher aperiodic intercept scores were associated with longer sleep onset latencies (β=2.36, *p*=0.01).

**Conclusion:** This study provides proof-of-concept that aperiodic activity may be predictive of vigilance. Given its practicality, cost-effectiveness and lower demand on health staff and patient time, it is suggested that aperiodic activity as a test of vigilance testing offers not only greater diagnostic objectivity but benefits to already resource-limited healthcare systems.

**Brief Summary:** The current gold standard for vigilance testing is the maintenance of wakefulness test (MWT) which measures the ability of an individual to maintain wakefulness in soporific conditions. Despite its advantages, the MWT is resource intensive and time consuming which has led to the exploration of alternative markers of wakefulness.

Our study provides proof of concept that neural aperiodic activity may be a viable alternative or adjunct to current vigilance tests. Further, we describe the ease at which analysis of the aperiodic activity is performed and the potential advantages of its utility in clinical practice.

## Introduction

In clinical sleep medicine, vigilance testing is performed to assess occupational and driving safety in individuals with sleep disorders. The maintenance of wakefulness test (MWT) measures the ability of an individual to maintain wakefulness in soporific conditions and is routinely performed in clinical and research settings to assess vigilance (1). As high levels of sleepiness impair vigilance and the ability to maintain wakefulness, the MWT can also be used to infer functional impairment due to sleepiness (2). Despite its advantages the MWT is resource-intensive and time-consuming. These challenges have stimulated interest in alternate measures. Previous studies in animal models have identified the down-regulation of cortical activity as a potential biomarker of sleep (3) and, possibly, sleepiness (4). Therefore, an alternative but untested approach to assess vigilance and, conversely, sleepiness is to directly examine cortical activity. Recent findings in humans indicate that the aperiodic EEG slope reflects arousal state and tracks the transition from wakefulness to drowsiness and sleep, suggesting it may serve as an objective marker of impaired vigilance (5,6). This is evidenced by reduced high-frequency power, increased low-frequency power, and steeper aperiodic EEG slopes (1/ *f* activity) reflective of increased inhibitory and decreased excitatory neural activity (7). However, the utility of the aperiodic EEG slope as a practical, less resource-intensive alternative to behavioural vigilance tests such as the MWT remains to be systematically evaluated in clinical populations.

The limitations of the MWT have been well-described (2). It is labour and patient-intensive, involving four 40-minute wake trials with strict protocols, and requires a specialised laboratory environment with at least one skilled technologist in attendance (1). Further, the interpretation of the MWT and its normative values are debated. The 2005 American Academy of Sleep Medicine (AASM) practice parameters quote a mean (SD) sleep latency of 30.4 ± 11.2 minutes with the lower limit of normal (LLN) of eight minutes (8). By contrast and discrepant to 2005 AASM practice parameters, Banks et al. (9) published normative data for the MWT in which the mean (SD) sleep latency (MSL) was 36.9 ± 5.4 minutes and the LLN was 26.1 minutes. Thus, raising concerns whether the recommended LLN for MSL are clinically meaningful. Further, it is unclear if statistically normal mean sleep latencies in a laboratory reflect the ability of an individual to focus on real-world tasks such as driving. In driving simulator studies and based on MWT findings, sleepy (MSL <19 minutes) compared to alert (MSL > 34 minutes) patients with untreated obstructive sleep apnoea demonstrate significantly lower number of road deviations (0.90 versus 0.35) and sleepy/very sleep (MSL = 19 – 33/<19 minutes) compared to alert (MSL > 33 minutes) controls and/or patients with untreated obstructive sleep apnoea/hypersomnia of central origin had a higher number of line crossings (1.53/1.50 versus 0.30) (10,11). However, in both the latter studies and despite the significant group differences, MSL and driving impairment values were poorly correlated in the sleepy patient groups thus raising questions about the thresholds used to classify sleepiness. In a naturalistic study involving two-hours of monotonous highway driving, MWT has also been reported to correlate poorly with driving performance irrespective of the MSL cut-off (12). Due to the trial design, the MWT also suffers from a ceiling effect thereby affecting confidence in making extrapolations about performance in real-life situations where driving duration may exceed the 40min WMT limit. It is noted that the MWT limit was increased from 20 to 40 minutes by Banks et al. (9) and that the capacity to maintain sustained vigilance is relevant in patient subpopulations such as commercial drivers who routinely perform long-haul trips (13). Finally, the MWT is subject to the influences of psychological factors, such as motivation (14), where patients may be aware of the sleep latency impacts on stimulant prescription and/or vehicle licensing/occupational approvals. Taken together, the latter factors raise concerns as to the ecological validity of the MWT and the need for alternate measures of vigilance/sleepiness. A potential measure is the aperiodic activity of the EEG signal.

Wake and sleep states are classically described as distinct patterns of oscillatory activity on EEG, which are divided into canonical frequency bands (15). For example, stage 2 sleep is characterized by sigma (12–15 Hz) and theta (4-7 Hz) oscillations, slow-wave sleep is predominantly characterized by sigma, delta (0.5-3.5 Hz) and slow oscillation (0–1 Hz) activity and REM sleep is dominated by wake-like theta oscillations (15). These states are also characterised by transient EEG patterns, such as sleep spindles and k-complexes. While the oscillatory activity of EEG has been extensively studied, there exists a less examined background non-oscillatory component which is referred to as the aperiodic or “scale-free” activity (16). Recent studies suggest that the aperiodic EEG component may have clinical utility. On a power spectral density plot, the aperiodic component is seen as a descending straight line that observes the 1/ *f* power-law relationship where lower frequencies exhibit greater amplitudes (i.e., power) and higher frequencies show decreased amplitude (17,18). Aperiodic slope variations have been associated with ageing and age-related cognitive decline (19), higher cognition and learning (20), cognitive speed (21), visuomotor learning and performance (22), schizophrenia (23), and states of arousal including sleep and anaesthesia (5).

It is presumed that sleepiness results from the accumulation of sleep need, known as Process S, although the neural marker for this process and how it can be measured on EEG has not been previously determined. The aperiodic slope, which represents the ratio of excitatory to inhibitory cortical activity captured by EEG, may be the breakthrough neural marker that can help determine sleep need. The aperiodic slope has been previously described in further detail by Chatburn, Lushington & Cross (24). We hypothesise that the aperiodic slope gradient may be a viable objective biomarker of sleep need and vigilance and could be used as an alternative or adjunct to the MWT (see Figure 1).

**Figure 1:**
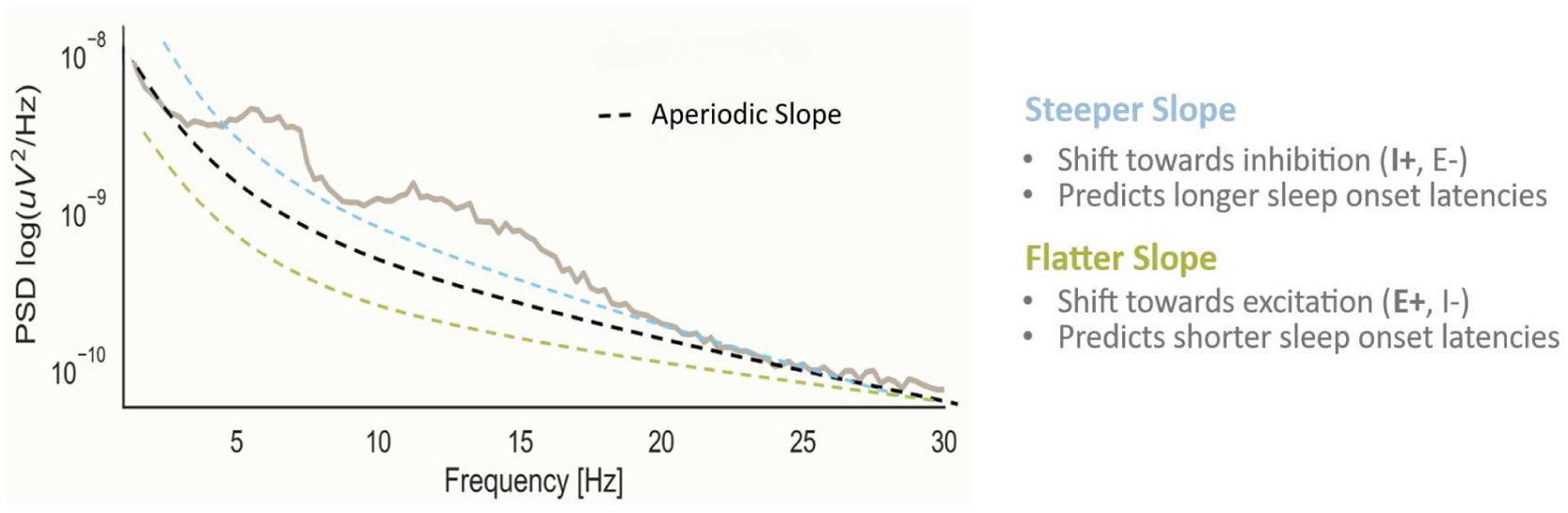
Illustration of the aperiodic slope and its hypothesized relationship to sleep onset latency^a^ The graph plots the log-transformed Power Spectral Density (PSD) of an EEG signal against Frequency (Hz). A steeper slope (dashed blue line) indicates a shift towards inhibition (I+, E-) and is associated with longer sleep onset latencies. Conversely, a flatter slope (dashed green line) indicates a shift towards excitation (E+, I-) and predicts shorter sleep onset latencies. **^a^**Figure adapted from Dziego et al., 2024 (34)

In sum, the aim of the present study is to test the relationship between resting state neural aperiodic activity and the ability of an individual to maintain wakefulness as assessed by MWT.

## Methods

### Participants

This is a retrospective study involving 50 patients referred from sleep medicine clinics to the Central Adelaide Local Health Network in South Australia between January 2009 and December 2023 for MWT to assess excessive daytime somnolence. As necessary, patient data was collected from review of electronic medical records and paper notes. All participants were included irrespective of medical comorbidities and medications given the real-world nature of the project. Self-rated sleep propensity was assessed at intake using the Epworth Sleepiness Scale (Mean (SD) = 4.54 (5.60), range 0-20, normal range 0-10), and regardless of score all participants were retained in the final sample. Ethics approval was obtained from Central Adelaide Local Health Network ethics committee (# 19734).

### Procedure

All participants undertook the MWT in accordance with guidelines published by the American Academy of Sleep Medicine (25). Four MWT trials were performed in a single workday, in which the participant was instructed to sit still with minimal movement and to remain awake for as long as possible. Trials were performed at two-hourly intervals, with standard MWT EEG *in situ*.

#### EEG

EEG was recorded across all MWT trials using either Compumedics E-series or Grael amplifier (Compumedics, Melbourne, Australia). Data were recorded at a sampling rate of 250Hz, with a 50Hz notch filter, from electrodes located at F3, C3, C4, O1 and O2 according to the international 10-20 system. Recording montages included sub-mental EMG and EOG located 1cm diagonally from the out canthus of each eye, bisecting the pupil. Impedances were maintained ≥10kΩ.

### Resting state analysis

The aperiodic components of the EEG (slope and intercept) were derived from MWT EEG using the IRASA method (26), as implemented in YASA (27). The recording involved participants sitting quietly with eyes opened in a dimly-lit testing room. Two-minute samples of EEG data were taken, were referenced to linked mastoids, broadband filtered to 30-45Hz and down-sampled to 100Hz using inbuilt functions in MNE-Python. Aperiodic estimates were calculated for each channel, but given the absence of a specific priori hypotheses regarding regional differences in aperiodic EEG activity, estimates were averaged across all channels to provide a global measure while minimizing the risk of spurious spatial findings due to multiple comparisons.

### Statistical analysis

Data inclusive of aperiodic variables and MWT outcomes, were imported into R and analysed using linear mixed models with restricted maximum likelihood (REML) estimates using *LME4*. We attempted to model sleep onset latency in the MWT separately from both aperiodic slope and ESS scores using the formulae:

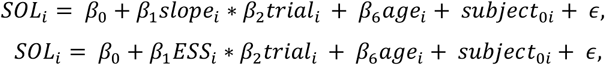

Where aperiodic slope (top model), ESS (bottom model) and trial are treated as fixed effects, age is treated as a covariate (due to the published effects of age on aperiodic EEG activity) and subject is treated as a random effect on the intercept, to account for between-subject effects on estimates. P-values were obtained using the LmerTest function and model outputs were extracted using Effects and plotted using GGplot2. Helmert contrasts were applied to trial, such that each trial was compared to the average of all preceding trials. Please note that all data visualisations use 85% confidence intervals and as such, non-overlapping confidence intervals indicate statistical significance at *p*<0.05.

## Results

### Patient demographics

Demographic data were available for all 50 participants (Table 1). The median age was 50 years and 66% were male. Most patients were overweight with median BMI 33. The ESS was low at the time of testing (median score 2) despite the test being performed for excessive somnolence. The most common sleep disorder was obstructive sleep apnoea in 14 of 18 patients with a documented sleep disorder. Comorbid depression was the most common psychiatric disorder. Patients were on a range of medications that may impact sleep, notably psychotropic medications such as antidepressants, stimulants and opiates (see Table 1).

**Table 1:**
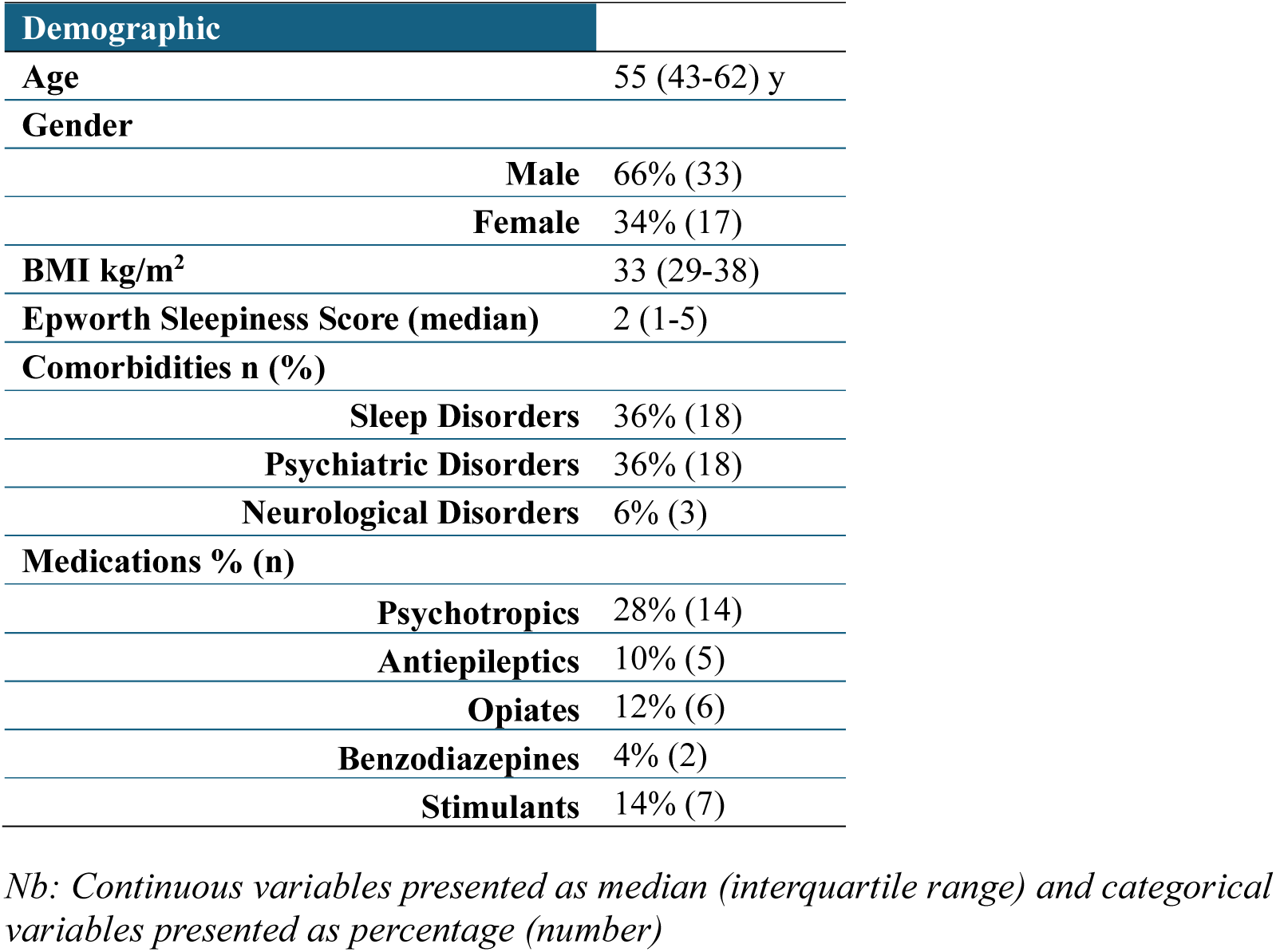
Patient Demographics (n=50)

| Demographic |  |
| --- | --- |
| Age | 55 (43-62) y |
| Gender |  |
| Male | 66% (33) |
| Female | 34% (17) |
| BMI kg/m<sup>2</sup> | 33 (29-38) |
| Epworth Sleepiness Score (median) | 2 (1-5) |
| Comorbidities n (%) |  |
| Sleep Disorders | 36% (18) |
| Psychiatric Disorders | 36% (18) |
| Neurological Disorders | 6% (3) |
| Medications % (n) |  |
| Psychotropics | 28% (14) |
| Antiepileptics | 10% (5) |
| Opiates | 12% (6) |
| Benzodiazepines | 4% (2) |
| Stimulants | 14% (7) |

*Nb: Continuous variables presented as median (interquartile range) and categorical variables presented as percentage (number)*

### Maintenance of wakefulness tests

The overall mean sleep latency (MSL) for all participants was 33.4 minutes. The median sleep latency for all participants was 40 (IQR 28-40). Most trials (72%) had reached the 40-minute ceiling. Only 1 participant satisfied criteria for excessive sleepiness based on an 8-minute LLN of MSL, whereas 11 reached criteria based on a LLN value of 26.1 minutes.

### Aperiodic slope is predictive of MWT SOL

A linear mixed model predicting sleep onset latency from aperiodic slope and MWT trial, while covarying for the effects of age and allowing random effects per participant, found that aperiodic slope was predictive of sleep onset latency in our sample population (β=-8.08, p=0.002). We did not observe a significant effect of trial (*p*=0.10) or significant interaction effects between slope and trial (*p*=0.18), suggesting that circadian effects did not reduce the predictive capacity of the aperiodic slope.

### ESS is not predictive of MWT SOL

To contextualise the previous result, and as an additional, exploratory analysis, we modelled sleep onset latency in the MWT using the ESS scores in the same manner as in the previous model. This model revealed no significant predictive capacity of the ESS in determining sleep onset latency in the MWT (β=-0.69, *p*=0.11). As before, we did not note a significant effect of trial, nor any significant interaction effects between trial and ESS.

### The aperiodic intercept predicts MWT SOL

As an exploratory analysis, we analysed the predictive capacity of the aperiodic intercept with a second linear mixed model predicting sleep onset latency, applying the same equation as used in previous analyses, with the same fixed/random/covariance arguments. Here, we found that the aperiodic intercept was predictive of sleep onset latency in our sample population (β=2.36, p=0.02). We did not observe a significant effect of trial (*p*=0.51), nor did we observe significant interaction effects between the intercept and trial (*p*=0.4). This relationship is depicted in figure 2. Comparing our intercept and slope models revealed no significant difference between them in terms of variance explained, although the estimated predicted error using the Akaike Information Criterion AIC for the slope model was slightly lower than that of the intercept model (slope model AIC = 1181.5; intercept model AIC = 1189.2).

**Figure 2:**
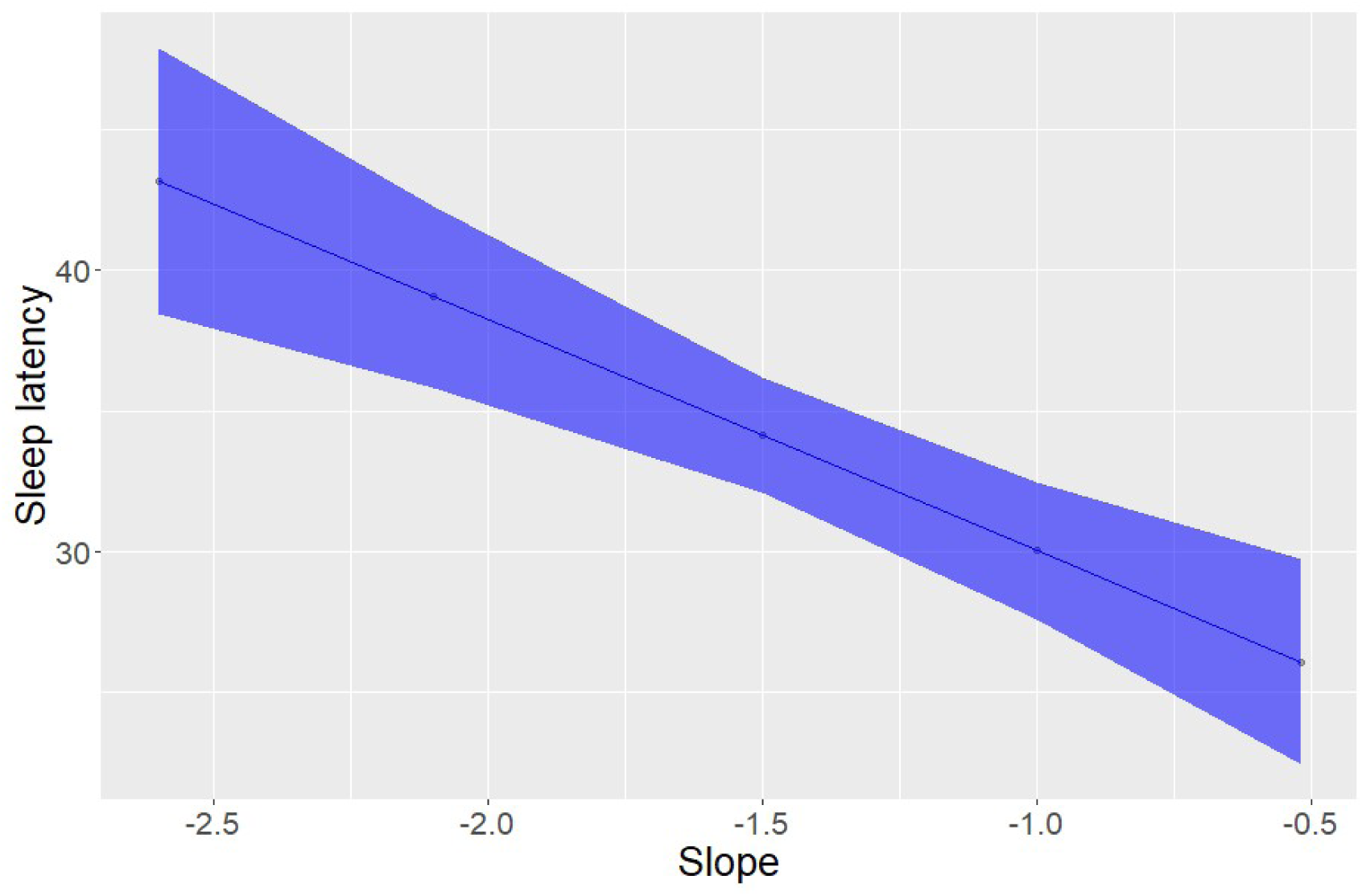
Linear mixed models predicting sleep onset latency from aperiodic slope *Aperiodic slope was predictive of sleep onset latency in the MWT, such that flatter slope indicated shorter sleep onset latency*.

**Figure 3:**
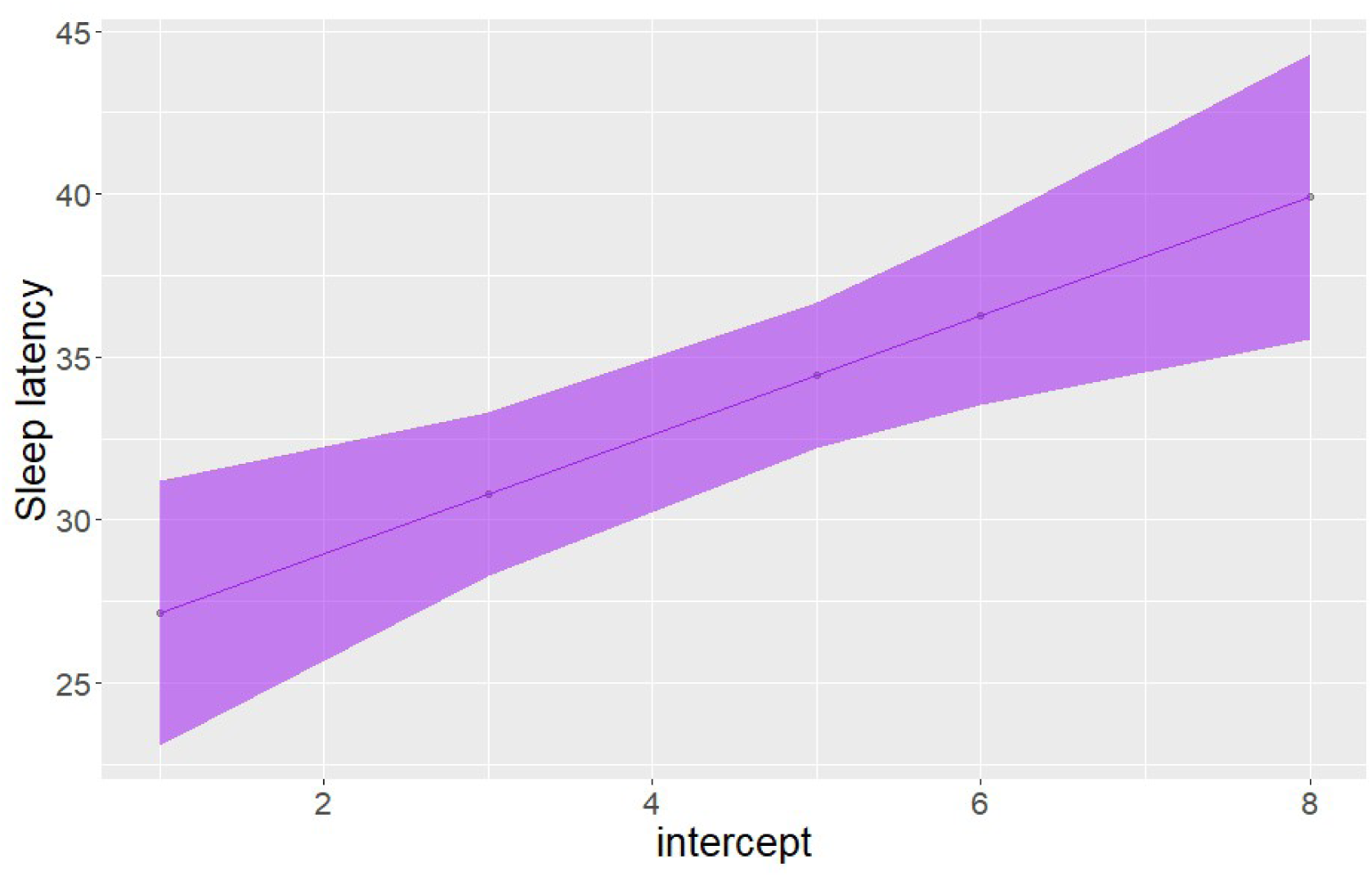
Linear mixed models predicting sleep onset latency from intercept scores *Correlation between intercept and sleep latency indicating that intercept scores are also related to sleep onset latency in the MWT*.

## Discussion

Our results demonstrate a potential clinical use of the aperiodic slope as an alternate diagnostic marker to the MWT and may provide insights into the neurobiology underlying normal and pathological sleepiness. Previous studies have demonstrated the ability of aperiodic slope in differentiating awake states, general anaesthesia, and NREM/REM sleep (5,28,29). Further, Gao & Richard (29) showed using computational models in animal studies that the excitation-inhibition ratio (E:I) of neuronal populations can be estimated from aperiodic slope such that steeper slopes were associated with increased inhibition, while other research (30–32) has demonstrated a shift in neuronal E:I balance with prolonged hours of wakefulness. Here, we have applied these findings to demonstrate a relationship between the awake state neural aperiodic slope and an individual’s ability to maintain vigilance as measured by MWT.

### Clinical application of aperiodic slope in the assessment of vigilance

Our study provides early evidence that steeper aperiodic slopes are predictive of longer sleep onset latencies. Given the limitations of the MWT, further study into this area is justified to further explore the role of aperiodic activity as a potential alternative or adjunct vigilance test. In real-world settings the advantages to this are multiple. Firstly, testing can be performed within minutes via standard EEG scalp electrodes obviating the need for prolonged, multiple 40-minute trials based on current labour-intensive MWT protocols. Secondly, patient tolerance and acceptance are likely to improve considering the shorter duration of testing, which in our study was two minutes of artefact-free EEG recording. Thirdly, the seemingly linear relationship between aperiodic slope and sleep latency shown in our study suggests that extrapolation of sleep onset latencies beyond 40 minutes may be possible, which could overcome the ceiling effect of the MWT and requires further investigation. Lastly, the aperiodic slope is obtained through objective short recordings of EEG data and cannot be easily manipulated, reducing the risk of biased outcomes due to (for example) motivational factors on the part of the patient.

Our exploratory analysis of the aperiodic intercept is of interest in understanding the basic neuronal operations which may be impacted by sleepiness. Broadly, while the aperiodic slope has been linked with the E:I ratio in neuronal populations, the aperiodic intercept may tag firing rates (33). Thus, our results suggest that there are multiple sequalae of clinical sleepiness in terms of the effects of the condition on the brain, the utility of which should be ascertained in future research. Further, our results may inform an ongoing discussion around the use of EEG in the objective measurement of sleepiness. While numerous oscillatory markers of sleepiness have been described in the literature (see Chatburn, Lushington & Cross, 2024 (24)), the development of these has generally been observational or atheoretical. As such, it is not clear exactly what is being measured when these are used. Our focus on the aperiodic EEG offers one solution to this, in that the aperiodic EEG has been linked to neuronal behaviours known to be influenced by increasing sleep need. Future research is needed to further elucidate the links between aperiodic EEG and sleep need in humans, including the best ways to measure the relationship and how to best use it in clinical and scientific applications.

### Limitations

There are numerous limitations in this study. Firstly, this data is from a small sample size of 50 participants and was collected retrospectively, which limits the generalisability of the findings.

Due to the retrospective design, all participants had a prior diagnosis of a sleep disorder, since MWTs are requested primarily for the evaluation of vigilance in patients with sleep disordered breathing and/or central disorders of hypersomnolence. Whilst this may have impacted the results, the MWT is commonly performed in a similar patient population and it can be argued that this sample reflects the likely target population of the technology, and thus affordance regarding ecological validity. Future works are needed to tease apart the effects of medication classes and clinical diagnoses on the predictive capacity of the aperiodic slope in the context of normal and clinical sleepiness. Finally, we observed a ceiling effect in that a majority of MWT trials achieved the maximum 40 minutes sleep latency as shown in Table 2. A possible explanation for this skew is that many participants would be highly motivated to maintain wakefulness given the potential occupational or legal implications of the test. There is also the possibility of bias in that clinicians are more likely to request an MWT once participants have been adequately treated for their sleep disorder. This was unavoidable considering the retrospective and real-world design of our study utilising the MWT as the current standard for vigilance testing which in a way reflects the practicalities of daily clinical practice.

### Implications for future research

The ideas presented herein can be further developed through a larger study involving multiple centres. A prospective trial design incorporating a control group of normal participants would improve generalisability and control confounding. Additionally, the real-world effect of aperiodic slope testing can be studied by coupling the test with a driving simulator as previously performed by Sagaspe et al. (10) for MWT to evaluate driving safety. Further, it may be informative for future research to link the pre-sleep aperiodic EEG with established markers of homeostatic sleep need (24). Doing so may allow for a better functional understanding of the neurophysiology of sleepiness and the mechanisms of dysfunction in central disorders of hypersomnolence.

## Conclusion

Our study is the first to demonstrate that the aperiodic EEG can predict sleep onset latency in the context of the MWT. Given the beneficial features of measuring aperiodic EEG activity including shorter-test time, reduced training and sleep laboratory resources to administer and greater tolerability of the test compared to MWT, clinical use is likely to be advantageous to a resource-limited healthcare system. Further research is needed to determine if the aperiodic EEG components are comparable to MWT and if it is capable of accurately determining vigilance in clinical practice.

## Data Availability

All data produced in the present study are available upon reasonable request to the authors, subject to ethics committee approval.

## Abbreviations

AASM: American Academy of Sleep Medicine
EEG: Electroencephalogram
EMG: Electromyography
EOG: Electrooculography
*f*: Frequency
IRASA: Irregular-Resampling Auto-Spectral Analysis
LLN: Lower limit of normal
MSL: Mean sleep latency
MWT: Maintenance of wakefulness test
REM: Rapid eye movement
SD: Standard deviation

## Acknowledgements

We would like to thank Ms Annaliese Anesbury for her assistance with the design and graphical rendering of Figure 1.

## Reference

1. Krahn LE, Arand DL, Avidan AY, Davila DG, DeBassio WA, Ruoff CM, et al. Recommended protocols for the Multiple Sleep Latency Test and Maintenance of Wakefulness Test in adults: guidance from the American Academy of Sleep Medicine. Journal of Clinical Sleep Medicine. 17(12):2489–98.

2. Taillard J, Micoulaud-Franchi JA, Martin VP, Peter-Derex L, Vecchierini MF. Objective evaluation of excessive daytime sleepiness. Neurophysiol Clin. 2024 Apr;54(2):102938.

3. Niethard N, Brodt S, Born J. Cell-Type-Specific Dynamics of Calcium Activity in Cortical Circuits over the Course of Slow-Wave Sleep and Rapid Eye Movement Sleep. J Neurosci. 2021 May 12;41(19):4212–22.

4. Vyazovskiy VV, Olcese U, Hanlon EC, Nir Y, Cirelli C, Tononi G. Local sleep in awake rats. Nature. 2011 Apr 28;472(7344):443–7.

5. Lendner JD, Helfrich RF, Mander BA, Romundstad L, Lin JJ, Walker MP, et al. An electrophysiological marker of arousal level in humans. Elife. 2020 July 28;9:e55092.

6. Nir Y, Andrillon T, Marmelshtein A, Suthana N, Cirelli C, Tononi G, et al. Selective neuronal lapses precede human cognitive lapses following sleep deprivation. Nat Med. 2017 Dec;23(12):1474–80.

7. Gerster M, Waterstraat G, Litvak V, Lehnertz K, Schnitzler A, Florin E, et al. Separating Neural Oscillations from Aperiodic 1/f Activity: Challenges and Recommendations. Neuroinformatics. 2022 Oct;20(4):991–1012.

8. Littner MR, Kushida C, Wise M, Davila DG, Morgenthaler T, Lee-Chiong T, et al. Practice parameters for clinical use of the multiple sleep latency test and the maintenance of wakefulness test. Sleep. 2005 Jan;28(1):113–21.

9. Banks S, Barnes M, Tarquinio N, Pierce RJ, Lack LC, McEvoy RD. The maintenance of wakefulness test in normal healthy subjects. Sleep. 2004 June 15;27(4):799–802.

10. Sagaspe P, Taillard J, Chaumet G, Guilleminault C, Coste O, Moore N, et al. Maintenance of wakefulness test as a predictor of driving performance in patients with untreated obstructive sleep apnea. Sleep. 2007 Mar;30(3):327–30.

11. Philip P, Chaufton C, Taillard J, Sagaspe P, Léger D, Raimondi M, et al. Maintenance of Wakefulness Test scores and driving performance in sleep disorder patients and controls. International Journal of Psychophysiology. 2013 Aug 1;89(2):195–202.

12. Virtanen I, Järvinen J, Anttalainen U. Can real-life driving ability be predicted by the Maintenance of Wakefulness Test? Traffic Injury Prevention. 2019 Aug 18;20(6):601–6.

13. Huhta R, Hirvonen K, Partinen M. Prevalence of sleep apnea and daytime sleepiness in professional truck drivers. Sleep Med. 2021 May;81:136–43.

14. Bonnet MH, Arand DL. Impact of motivation on Multiple Sleep Latency Test and Maintenance of Wakefulness Test measurements. J Clin Sleep Med. 2005 Oct 15;1(4):386–90.

15. Brown RE, Basheer R, McKenna JT, Strecker RE, McCarley RW. Control of sleep and wakefulness. Physiol Rev. 2012 July;92(3):1087–187.

16. Donoghue T, Haller M, Peterson EJ, Varma P, Sebastian P, Gao R, et al. Parameterizing neural power spectra into periodic and aperiodic components. Nat Neurosci. 2020 Dec;23(12):1655–65.

17. Miller KJ, Sorensen LB, Ojemann JG, den Nijs M. Power-law scaling in the brain surface electric potential. PLoS Comput Biol. 2009 Dec;5(12):e1000609.

18. He BJ. Scale-free brain activity: past, present, and future. Trends in Cognitive Sciences. 2014 Sept 1;18(9):480–7.

19. Voytek B, Kramer MA, Case J, Lepage KQ, Tempesta ZR, Knight RT, et al. Age-Related Changes in 1/f Neural Electrophysiological Noise. J Neurosci. 2015 Sept 23;35(38):13257–65.

20. Cross ZR, Corcoran AW, Schlesewsky M, Kohler MJ, Bornkessel-Schlesewsky I. Oscillatory and Aperiodic Neural Activity Jointly Predict Language Learning. J Cogn Neurosci. 2022 Aug 1;34(9):1630–49.

21. Ouyang G, Hildebrandt A, Schmitz F, Herrmann CS. Decomposing alpha and 1/f brain activities reveals their differential associations with cognitive processing speed. Neuroimage. 2020 Jan 15;205:116304.

22. Immink MA, Cross ZR, Chatburn A, Baumeister J, Schlesewsky M, Bornkessel-Schlesewsky I. Resting-state aperiodic neural dynamics predict individual differences in visuomotor performance and learning. Hum Mov Sci. 2021 Aug;78:102829.

23. Molina JL, Voytek B, Thomas ML, Joshi YB, Bhakta SG, Talledo JA, et al. Memantine Effects on Electroencephalographic Measures of Putative Excitatory/Inhibitory Balance in Schizophrenia. Biol Psychiatry Cogn Neurosci Neuroimaging. 2020 June;5(6):562–8.

24. Chatburn A, Lushington K, Cross ZR. Considerations towards a neurobiologically-informed EEG measurement of sleepiness. Brain Res. 2024 Oct 15;1841:149088.

25. Littner MR, Kushida C, Wise M, G. Davila D, Morgenthaler T, Lee-Chiong T, et al. Practice Parameters for Clinical Use of the Multiple Sleep Latency Test and the Maintenance of Wakefulness Test. Sleep. 2005 Jan 1;28(1):113–21.

26. Wen H, Liu Z. Separating Fractal and Oscillatory Components in the Power Spectrum of Neurophysiological Signal. Brain Topogr. 2016 Jan;29(1):13–26.

27. Vallat R, Walker MP. An open-source, high-performance tool for automated sleep staging. Elife. 2021 Oct 14;10:e70092.

28. Colombo MA, Napolitani M, Boly M, Gosseries O, Casarotto S, Rosanova M, et al. The spectral exponent of the resting EEG indexes the presence of consciousness during unresponsiveness induced by propofol, xenon, and ketamine. Neuroimage. 2019 Apr 1;189:631–44.

29. Gao R. Interpreting the electrophysiological power spectrum. J Neurophysiol. 2016 Feb 1;115(2):628–30.

30. Huber R, Mäki H, Rosanova M, Casarotto S, Canali P, Casali AG, et al. Human cortical excitability increases with time awake. Cereb Cortex. 2013 Feb;23(2):332–8.

31. Mroczek M, de Grado A, Pia H, Nochi Z, Tankisi H. Effects of sleep deprivation on cortical excitability: A threshold-tracking TMS study and review of the literature. Clin Neurophysiol Pract. 2024;9:13–20.

32. De Gennaro L, Marzano C, Veniero D, Moroni F, Fratello F, Curcio G, et al. Neurophysiological correlates of sleepiness: a combined TMS and EEG study. Neuroimage. 2007 July 15;36(4):1277–87.

33. Bai D, Hu J, Jülich S, Lei X. Impact of sleep deprivation on aperiodic activity: a resting-state EEG study. J Neurophysiol. 2024 Nov 1;132(5):1577–88.

34. Dziego CA, Bornkessel-Schlesewsky I, Schlesewsky M, Sinha R, Immink MA, Cross ZR. Augmenting complex and dynamic performance through mindfulness-based cognitive training: An evaluation of training adherence, trait mindfulness, personality and resting-state EEG. PLOS ONE. 2024 May 20;19(5):e0292501.

